# Evaluating the Performance of the DPP® ZDC IgM/IgG Rapid Test for Chikungunya Virus Diagnosis in Febrile Outpatients: A Prospective, Diagnostic Accuracy Study

**DOI:** 10.1101/2023.04.27.23289217

**Authors:** José Moreira, Janaina Barros, B. Leticia Fernandez-Carballo, Camille Escadafal, Guilherme S. Ribeiro, Sabine Dittrich, Patrícia Brasil, André M. Siqueira

## Abstract

**Objective:** Evaluate the performance of a novel antibody-based rapid diagnostic test (RDT) for detecting Chikungunya virus (CHIKV) infection in febrile patients in Rio de Janeiro, Brazil.

**Methods:** We prospectively enrolled non-severe febrile patients aged 2-65 years presenting as outpatients between October 2018 and July 2019. Serum samples were collected during acute and convalescent phases and tested for CHIKV antibodies using the DPP® ZDC IgM/IgG rapid test and compared against the reference test, CHIKV RT-PCR. We determined the seropositivity using ELISA IgM/IgG and evaluated the diagnostic performance of the WHO-endorsed CHIKV clinical definition against the reference test.

**Results:** Of 500 participants, 226/261 (86.5%) tested ELISA IgM positive, 45/271 (16.6%) tested ELISA IgG positive, 100/294 (34%) CHIKV RT-PCR positive, and 117/495 (23.6%) RDT-antibody positive. During the acute phase [median 3 (2-4) days post illness onset], the sensitivity of IgM, IgG, and combined IgM/IgG ranged from 14.71-34.85%, while specificity ranged from 63.32-65.61%. During the convalescent phase [mean 16.5 (±5.5) days post-illness onset], sensitivity increased from 65.75% to 77.78%, and specificity ranged from 93.33-98.11%. The WHO’s CHIKV clinical definition had a sensitivity, specificity, positive predictive value, and negative predictive value of 88 (79.9-93.6)%, 74 (68-80)%, 64.2 (58.2-69.8)%, and 92.3 (87.6-95.3)%, respectively.

**Conclusions:** The DPP® ZDC IgM/IgG accurately diagnosed CHIKV on samples collected during the convalescent phase. Field applications include investigating CHIKV in patients with sub-acute to chronic osteoarticular symptoms and conducting serosurveys to inform priority areas for CHIKV vaccine implementation. The WHO’s clinical definition of CHIKV was accurate and could be deployed, especially in regions with limited diagnostic capacity.

**Clinicaltrials.gov:** NCT03047642

## Introduction

The Chikungunya virus (CHIKV) is a re-emerging mosquito-borne alphavirus posing a significant public health threat in tropical and subtropical regions of the globe. Infection caused by CHIKV is characterized by an acute onset of high-grade fever, rash, and arthralgia [1]. The first autochthonous cases of CHIKV in Brazil were confirmed in cities located in the north and northeast part of the country in 2014 [2]. Since then, an explosive epidemic has emerged, and more than 930.000 confirmed cases have been notified over the seven years [3]. Unfortunately, in most parts of Brazil, the paucity of laboratories with CHIKV molecular testing is restricting case detection and outbreak management. Furthermore, reverse transcriptase-polymerase chain reaction (RT-PCR) testing is costly, requires well-equipped laboratories and well-trained personnel. Thus, there is a clear need to develop new strategies for CHIKV testing and outbreak detection that do not rely entirely on RT-PCR. CHIKV antibody-based rapid diagnostic tests (RDTs) are becoming available to meet this need, but little information exists regarding their performance in samples originated from Latin America [4-6]. Thus, we conducted a prospective study of febrile patients to evaluate the performance characteristics of a novel antibody-based CHIKV RDT compared with gold-standard CHIKV RT-PCR. The primary outcomes were to (i) describe the positivity rate by CHIKV diagnostic assay tested [RDT, RT-PCR and Enzyme-linked Immunosorbent assay (ELISA)].; (ii) describe the CHIKV-infected population and compare it to uninfected patients; (iii) evaluate the performance of the RDT against the gold standard during the acute and convalescent-phase; (iv) compare the agreement of RDT against the ELISA assays, and (v) examine the performance of the current CHIKV clinical definition endorsed by the WHO against the reference test.

## Methods

### Study design and participants

This is a *post-hoc* analysis of the Biomarker for Fever-Diagnostic (BFF-Dx) study enrolling non-severe febrile participants in three low-and middle-income countries, aiming to evaluate biomarkers to distinguish bacterial from non-bacterial causes of fever [7]. The Brazilian National Ethics Research Committee approved the study (IRB# 70984617.9.0000.5262), and written informed consent was obtained from all participants or the caregivers of participants before enrollment. We reported as per the Standards for Reporting Diagnostic Accuracy (STARD) guideline (see Supplementary appendix).

A prospective observational study was conducted for consecutive patients 2-65 years presenting with an axillary temperature ≥ 37.5° C for a week or less or those with a recent fever history. According to triage protocols, those who had signs of severe disease and those who were unlikely to follow the study procedures were excluded and advised to continue receiving the clinic’s routine care service. We screened for febrile participants at two primary care clinics and two emergency departments in Rio de Janeiro, Brazil, between October 28^th^, 2018, to July 31^st^, 2019, at a time when a large CHIKV outbreak was taking place. The state surveillance data has registered an increase of almost 300% of the CHIKV cases reported in 2019 compared to the same period in the previous year [8].

We performed a detailed clinical and microbiological investigation to identify the cause of fever by performing a large panel of laboratory tests based on a symptom-driven approach [7].

Acute and convalescent-blood samples of those with undifferentiated febrile illness (i.e., without fever focus on clinical examination and history taking) were drawn for arboviral diagnosis. Samples were refrigerated until centrifugation and obtained sera were stored at – 80°C for serological and molecular testing.

### Detection of CHIKV RNA with RT-PCR testing

Acute sera from acute undifferentiated febrile patients were submitted to RNA extraction and tested by RT-PCR for CHIKV. Briefly, viral RNA was extracted using the QIAmp Viral RNA isolation kit (QIAGEN, Hilden, Germany). Subsequently, RT-PCR (Altona, Astra Diagnostics, Hamburg, Germany) was performed on the extraction product using specific primers to identify CHIKV RNA [9]. We categorized viral loads according to the Ct values obtained at the initial medical visit: high viral load group (i.e., ct <25) and a low viral load group (i.e., ct ≥25)

### Detection of CHIKV-specific IgM and IgG antibodies by ELISA

Acute serum samples were assayed for the presence of IgM and IgG antibodies against CHIKV using the anti-Chikungunya IgM virus ELISA test (Euroimmun, Luebeck, Germany) and the anti-Chikungunya IgG virus ELISA test kit (Euroimmun, Luebeck, Germany) as previously described [10]. A manual microplate reader performed the ELISA plate optical density reading. The optical density ratio obtained from the patients’ serum and the calibrator were interpreted according to the manufacturer’s instructions. Samples yielding equivocal/borderline ratio results were repeated once, and second results were considered final.

### CHIKV antibody-based RDT

The DPP^®^ ZDC IgM/IgG (Bio-Manguinhos, Fundação Oswaldo Cruz, Brazil) is a newly developed automated dual-path immunochromatographic platform for Zika, Chikungunya, and Dengue infections. The kit results from a partnership between Bio-Manguinhos (Fundação Oswaldo Cruz, Rio de Janeiro, Brazil) and the Chembio Diagnostic, Inc., USA, and uses whole blood or serum samples for the simultaneous detection of IgM and IgG against the three most widely circulating arboviruses in Brazil. For this study, we evaluated the performance of this assay using frozen serum samples collected from acute and convalescent phase of the disease. The Brazilian regulatory agency-ANVISA - has approved (registration no. 80142170035) the clinical use of the study’s RDT kits to perform the differential diagnosis of the three arboviruses at the bedside. Results become available in 15-20 minutes, and a microreader was then used to provide an unbiased test interpretation (i.e., reported as reactive, non-reactive, or undetermined). The cut-off values for reactive, non-reactive and indetermined were ≥ 22, ≤ 18, and > 18 and < 22, respectively. Indetermined results were dismissed and not take into analysis. A trained operator performed the RDT according to the manufacturer’s instructions and read the results in the laboratory, blinded to the results of the other CHIKV assays. The RDT was stored at room temperature (<30°C) before testing.

### CHIKV clinical case definition

We then examined the performance of the CHIKV clinical case definition endorsed by the World Health Organization (WHO) [11] against the gold-standard RT-PCR, which can be especially useful in the context of an outbreak, where sufficient time or resources for compulsory laboratory confirmation is not practical.

### Statistical analysis

Descriptive statistics summarized the characteristics and distribution of values in the cohort [mean ± standard deviation or median and interquartile range], and the Shapiro-Wilk test was used to assess normality. The diagnostic performance parameters (i.e., sensibility, specificity, positive predicted value, negative predictive value, and the area under the curve) and their respective 95% confidence intervals (CI) of the RDT were estimated and compared to the CHIKV RT-PCR results [12]. Similarly, the performance of the WHO CHIKV clinical definition was evaluated against the RT-PCR results. The CIs were estimated using the Wilson method. The inter-rater agreement between RDT and the ELISA assays was determined, and the corresponding kappa scores were calculated. The strength of agreement was interpreted using the Landis and Koch criteria [13].

## Results

During the study period, 500 participants were enrolled. Figure 1 shows the laboratory flowchart for the study population. The diagnostic positivity rate for CHIKV was much higher with IgM-based ELISA alone (138/261, 52.8%), followed by a combination of RT-PCR and IgM-based ELISA (29/162,17.9%), and RT-PCR alone (44/294,14.9%) (see Supplementary Table 1). Supplementary Figure 1 depicts the positivity across different diagnostic assays according to the time elapsed between symptom onset and sample collection. Detection of the CHIKV RT-PCR persisted for an extended period, up to 10 days after the illness (Supplementary Figure 2); however, in 81% of the cases, CHIKV RNA was detected in samples drawn up to three days of illness. Detection of IgM by ELISA occurred up to 10 days after the illness, but most positive results (72.5%) were detected in samples drawn within the three days of illness. Finally, detection of IgG by ELISA occurred up to 7 days after the illness, and more than half of the samples (57.7%) were positive after three days of illness.

**Figure 1.**
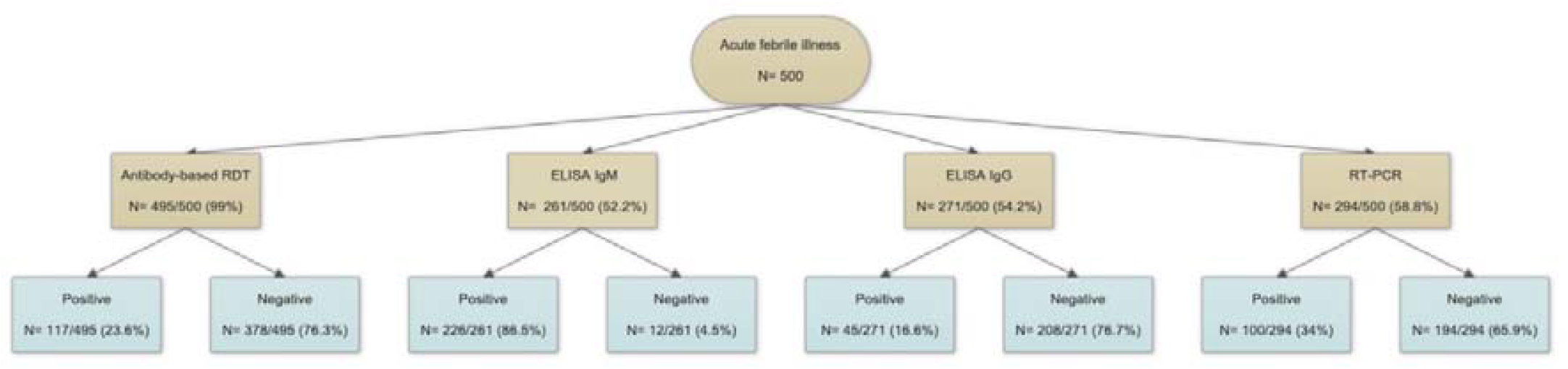
Laboratory flowchart of the study population, Rio de Janeiro, Brazil, October 2018-July 2019 ELISA: Enzyme-linked immunosorbent assay; RT-PCR: Reverse transcriptase polymerase chain reaction; RDT: Rapid diagnostic test; IgM: Immunoglobulin M; IgG: Immunoglobulin G.

Supplementary Figure 3 shows the viral kinetics (measured in Ct values) according to the time after illness onset. The odds of presenting a lower Ct value were 35.5 times higher in those that sought care within three days after illness onset than those who sought care later in the disease course (OR: 35.5 (8.71-144.72)].

We confirmed CHIKV infection in 100/294 (34%) patients (i.e., CHIKV was confirmed by a positive sample according to CHIKV RT-PCR). Supplementary Table 2 shows the baseline characteristics according to CHIKV infection. Patients with confirmed CHIKV infection were more likely to be males (unadjusted OR: 1.96 [1.20-3.22]), older (unadjusted OR: 1.02 [1.01-1.04]), and presented early after the onset of symptoms (unadjusted OR: 0.77 [0.65-0.90]) compared to CHIKV-negative patients. The most commonly observed symptoms among those with confirmed CHIKV infection were headache (90%), arthralgia (88%), rash (54%), redness of the eyes (38%), and photophobia (38%). Regarding the laboratory abnormalities, confirmed CHIKV-infected patients presented predominately with lymphocytopenia (76%), and more than half had serum C-reactive protein levels ≥ 5mg/dl (59%).

During the acute-phase samples [median 3 (2-4) days post illness onset], the performance of the RDT IgM and IgG individually, and both combined, is shown in Table 1. Overall, RDT IgM, IgG, and the analysis combining both of them revealed poor sensitivity (range: 14.71-34.85%) and modest specificity (range: 63.32-65.61%).

**Table 1.** Diagnostic performance of the acute-phase febrile samples for IgM, IgG and the combination of both IgM and IgG components of the DPP® ZDC IgM/IgG test (Bio-Manguinhos, Fiocruz, Brazil) against Chikungunya RT-PCR, among non-severe febrile patients, Rio de Janeiro, Brazil, October 2018-July 2019.

| Index test | N,<br>sensitivity<br>(95% CI) | N,<br>specificity<br>(95% CI) | PPV<br>(95% CI) | NPV<br>(95% CI) | PLR<br>(95%<br>CI) | NLR<br>(95%<br>CI) | AUC<br>(95%<br>CI) |
| --- | --- | --- | --- | --- | --- | --- | --- |
| **DPP IgM** | 34.85 (23.53-47.58) | 65.61 (58.94-71.85) | 23.23 (15.33-32.79) | 77.13 (70.45-82.93) | 1.01 (0.70-1.48) | 0.99 (0.81-1.21) | 58.54 (52.60-64.30) |
| **DPP IgG** | 14.71 (4.95-31.06) | 63.32 (57.13-69.20) | 5.00 (1.64-11.28) | 84.97 (79.14-89.70) | 0.40 (0.18-0.92) | 1.35 (1.14-1.59) | 57.68 (51.80-63.40) |
| **DPP IgM/IgG** | 32.39 (21.76-44.55) | 63.77 (57.67-69.57) | 19.33 (12.66-27.58) | 77.88 (71.76-83.22) | 0.89 (0.62-1.30) | 1.06 (0.88-1.28) | 57.14 (51.66-62.50) |

AUC stands for area under the curve; CI confidence intervals; NPV negative predictive value; NLR negative likelihood ratio; PPV positive predictive value; PLR positive likelihood ratio

During the convalescent-phase samples [mean 16.5 (±5.5) days post illness onset], the sensitivity of the IgM and IgG increased to 65.75% and 77.78%, while the specificity increased to 98.11% and 93.33%, respectively (Supplementary Table 3).

We then compared the agreement of the RDT results with the ELISA results. The RDT IgM showed a slight agreement with ELISA IgM (24 % concordance, k coefficient = 0.01; *n* = 56), whereas the RDT IgG had a substantial agreement with ELISA IgG (92.8% concordance, k coefficient = 0.70; *n*= 233).

Supplementary Figure 4 depicts the diagnostic accuracy of the current CHIKV’s clinical definition endorsed by the WHO compared to RT-PCR. The sensibility, specificity, positive predictive value, and negative predictive value was 88 (79.9-93.6) %, 74 (68-80) %, 64.2 (58.2-69.8) %, and 92.3 (87.6-95.3) %, respectively.

## Discussion

This study presents a clinical evaluation of a newly developed antibody-based RDT compared to the gold-standard CHIKV RT-PCR in a cohort of non-severely ill febrile patients in Rio de Janeiro, Brazil during a large CHIKV outbreak. This novel assay aimed to detect CHIKV antibodies and address a knowledge gap regarding RDT performance in Latin America samples, where RT-PCR is not sufficiently accessible. We found an overall poor sensitivity and modest specificity during the acute-phase samples, while the sensitivity and specificity increased during the convalescent-phase samples, corroborating with CHIKV antibody dynamics. Additionally, the study found a slight agreement between the RDT IgM and ELISA IgM results, whereas the RDT IgG showed a substantial agreement with ELISA IgG. Finally, the diagnostic accuracy of the WHO’s CHIKV clinical definition demonstrated good accuracy compared to RT-PCR, being a potential solution during outbreak where laboratory confirmation of all affected individuals is impractical.

A significant strength of our study is its prospective design, enabling assessment of RDT performance in a real-world clinical setting. Additionally, the study was conducted during a large CHIKV outbreak, enhancing the external validity. Our study adds valuable information on the dynamics of different CHIKV assays (RDT, ELISA and RT-PCR) throughout the disease course, shedding light on the persistence of IgM in regions with CHIKV circulation. For instance, more than 70% of samples were positive for ELISA-IgM within the first three days post-onset of illness, potentially suggesting past infection rather than recent infection. However, some limitations should be acknowledged. First, the RDT performance was evaluated using frozen serum samples rather than fresh samples, which might have affected its sensitivity and specificity. Evaluating test performance at bedside using capillary blood samples and after the recommended reading time should be further evaluated. Second, the study population’s age range was limited to 2-65 years, which might not fully represent RDT performance in younger or older populations. Third, we could not perform molecular and serological CHIKV testing for the entire cohort, limiting comprehensive comparative analysis between the assays. This is because the volume of samples was firstly dedicated to the primary analysis of the BFF-Dx study (i.e., standard of care testing and host-based biomarkers of infection). Finally, the RDT performance was evaluated during a time when the East-Central-South-African (ECSA) genotype predominated. Others have found different sensitivity of RDT assays when analyzing separate outbreaks caused by different CHIVK variants [14]. A previous genomic study revealed that the ECSA genotype of CHIKV had been circulating undetected in Rio long before notification by the state surveillance department [15], while information about the genotype used in the RDT assay evaluated is unknown. Therefore, more evidence is needed to clarify the CHIKV antibody assay performance in samples collected from distinct regions where different CHIKV lineages circulate.

Our data corroborates recent findings of a systematic review of serologic tests for CHIKV infection diagnosis [16]. This review found that IgM and IgG antibody detection tests had high accuracy (>90%) for samples collected in the convalescent phase of CHIKV infection. Conversely, the sensitivity of the IgM detection tests was low for acute-phase samples (26.2%) compared to the convalescent-phase samples (98.4%). Thus, the findings of the review, along with our findings, confirm that serologic assay are not recommended for samples taken during the acute phase of infection, as early in the disease, antibodies levels can be below the detection limit of most serological assays. We suggest adopting the DPP® ZDC IgM/IgG test (Bio-Manguinhos, Fiocruz, Brazil) to investigate patients with sub-acute and chronic osteomuscular symptoms and in serosurveys to inform the priority regions for potential CHIKV vaccine introduction in endemic countries.

We have previously advocated for the need to evaluate the feasibility, acceptability, cost-effectiveness, sustainability, and policy testing issues before widespread adoption of CHIKV RDTs in endemic countries [17]. Furthermore, it is imperative that we assess the impact of CHIKV RDTs on integrated fever case management and how their implementation translates into better prescription practices for acute febrile patients (i.e.; reducing unnecessary antibiotic prescription and increasing referrals for patients with osteoarticular symptoms to rheumatological and physiotherapeutic services). Chikungunya-endemic countries should prioritize the development of a policy testing strategy for identifying the patient population that would benefit the most from RDT and determining how to manage patients according to the RDT results.

To conclude, this study sheds light on the performance of a novel antibody-based RDT in the context of a large CHIKV outbreak and highlights the challenges associated with diagnosing CHIKV infections in endemic settings. The results emphasize the need for further advancements in diagnostic tools to enhance their accuracy and applicability in diverse populations and different stages of the disease. As chikungunya-endemic countries continue to grapple with the public health challenges posed by this disease, it is crucial to prioritize the development and evaluation of new diagnostic tools, while also optimizing the use of existing ones. A multidisciplinary approach that integrates clinical, serological, and molecular diagnostic methods, as well as robust policy testing strategies, will be key to improving patient management and health outcomes in the face of ongoing CHIKV outbreaks.

## Data Availability

All data produced in the present work are contained in the manuscript

## Declarations

### Ethics approval and consent to participate

The Brazilian National Ethics Research Committee approved the study (IRB# 70984617.9.0000.5262), and written informed consent was obtained from all participants or the caregivers of participants before enrollment.

### Consent for publication

Not applicable.

### Availability of data and materials

All data generated or analyzed during this study are included in this published article and its supplementary information files.

### Competing interests

JM is an employee of Instituto Butantan (Sao Paulo, Brazil). SD, LC, CE are employees or formal employees of Foundation for Innovative New Diagnostics (FIND).

### Funding

This work was supported by independent grants from Australia, the Netherlands, and the UK aid from the British people. The funding bodies did not play any role in the study’s design and collection, analysis, interpretation of data, or manuscript writing.

### Authors’ contributions

JM was responsible for design the experiment, data acquisition, analysis, interpretation and drafted the work; JB design the work and did the laboratory analysis and interpretation; LC, CE and SD were responsible for data interpretation and revision of the draft; GR made substantial contributions on the conception and interpretation of data, PB and AS revised the final draft prior to submission. All authors critically reviewed the manuscript, gave their final approvals, and are accountable for accuracy and integrity.

### Authors’ information

JM is a physician-scientist specializing in tropical medicine and currently serves as the Clinical Development Medical Director at Instituto Butantan. His works includes the epidemiology of acute febrile illness in Latin America, the identification of host biomarkers to differentiate bacterial from non-bacterial infections, and the clinical development of randomized trials with experimental arbovirus vaccines to evaluate their efficacy, safety and immunogenicity.

## Acknowledgments

The authors want to thank all participants and their family members who have contributed to these efforts. The authors also want to acknowledge the help of all of the clinical, laboratory, and administrative staff at the UPA Manguinhos, UPA Rocha Miranda, Centro de Saude Escola Germano Sinval Faria (CSEGSF), and Clinica da Familia Armando Palhares Aguinaga (CFAPA). This study would not have been possible without their support. JM received a scholarship from Fundacao de Amparo a Pesquisa do Estado do Rio de Janeiro (FAPERJ) for his Ph.D. study in Brazil.

## Appendix Table of Contents

This supplementary material has been provided by the authors to give readers additional information about their work.

## Supplementary Tables

**Supplementary Table 1.** Number of positive Chikungunya diagnostic assays tested among non-severe febrile patients, Rio de Janeiro, Brazil, October 2018-July 2019

| CHIKV assay |  |
| --- | --- |
| ELISA IgM POSITIVE | 138 |
| RT-PCR POSITIVE | 44 |
| RT-PCR POSITIVE/ ELISA IgM POSITIVE | 29 |
| RDT IgM POSITIVE | 25 |
| RDT IgM POSITIVE/RDT IgG POSITIVE | 21 |
| RDT IgM POSITIVE/RDT IgG POSITIVE/ ELISA IgM POSITIVE/ ELISA IgG POSITIVE | 17 |
| RDT IgM POSITIVE/ ELISA IgM POSITIVE | 14 |
| RT-PCR POSITIVE/RDT IgM POSITIVE | 12 |
| ELISA IgM POSITIVE/ ELISA IgG POSITIVE | 8 |
| RDT IgG POSITIVE | 6 |
| RT-PCR POSITIVE/RDT IgM POSITIVE/ ELISA IgM POSITIVE | 5 |
| RDT IgG POSITIVE/ ELISA IgM POSITIVE/ ELISA IgG POSITIVE | 4 |
| RT-PCR POSITIVE/RDT IgM POSITIVE/RDT IgG POSITIVE/ ELISA IgM POSITIVE/ ELISA IgG POSITIVE | 3 |
| RDT IgM POSITIVE/ ELISA IgM POSITIVE/ ELISA IgG POSITIVE | 3 |
| RT-PCR POSITIVE/RDT IgM POSITIVE/ ELISA IgM POSITIVE/ ELISA IgG POSITIVE | 2 |
| RT-PCR POSITIVE/ ELISA IgM POSITIVE/ ELISA IgG POSITIVE | 2 |
| RDT IgM POSITIVE/RDT IgG POSITIVE/ ELISA IgG POSITIVE | 2 |
| ELISA IgG POSITIVE | 2 |
| RT-PCR POSITIVE/RDT IgM POSITIVE/RDT IgG POSITIVE/ ELISA IgM POSITIVE | 1 |
| RT-PCR POSITIVE/RDT IgG POSITIVE | 1 |
| RT-PCR POSITIVE/ ELISA IgG POSITIVE | 1 |
| RDT IgM POSITIVE/ ELISA IgG POSITIVE | 1 |

We applied a combination of molecular and serologic diagnostic assays to evaluate the diagnosis of Chikungunya (CHIKV) in our cohort. ELISA IgM alone had the highest positivity rate (138/261, 52.8%).

**Supplementary Table 2.**
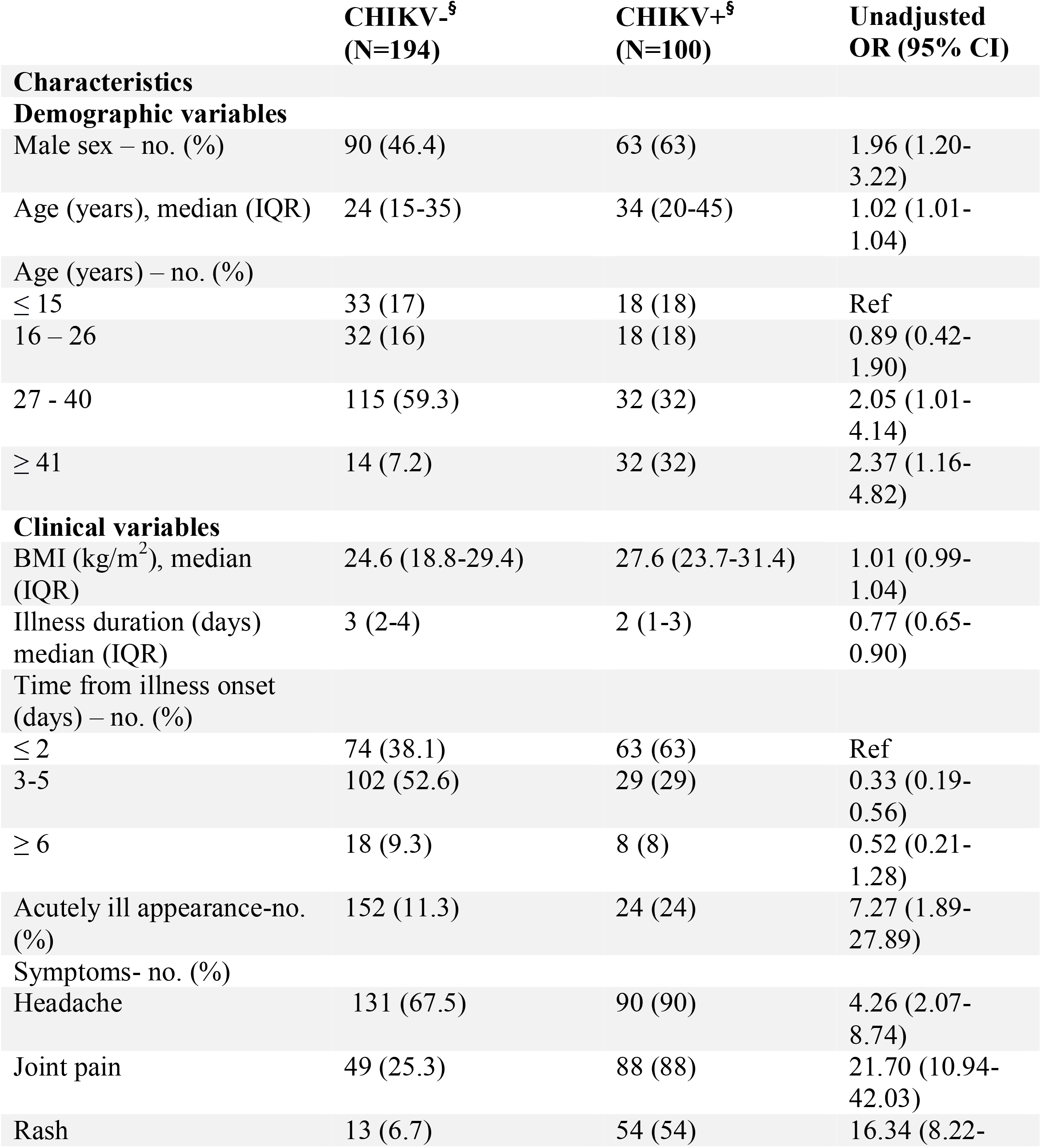
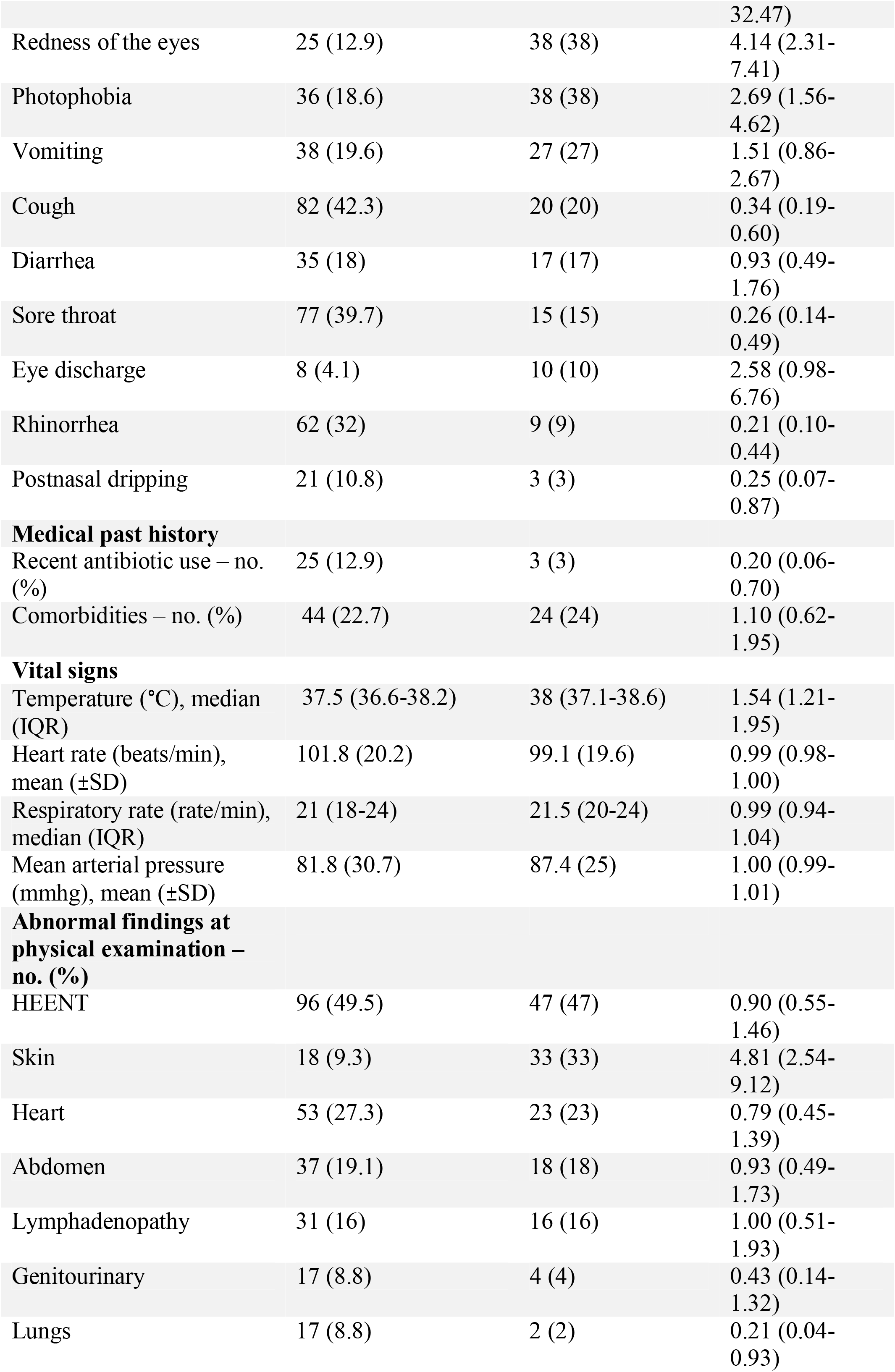
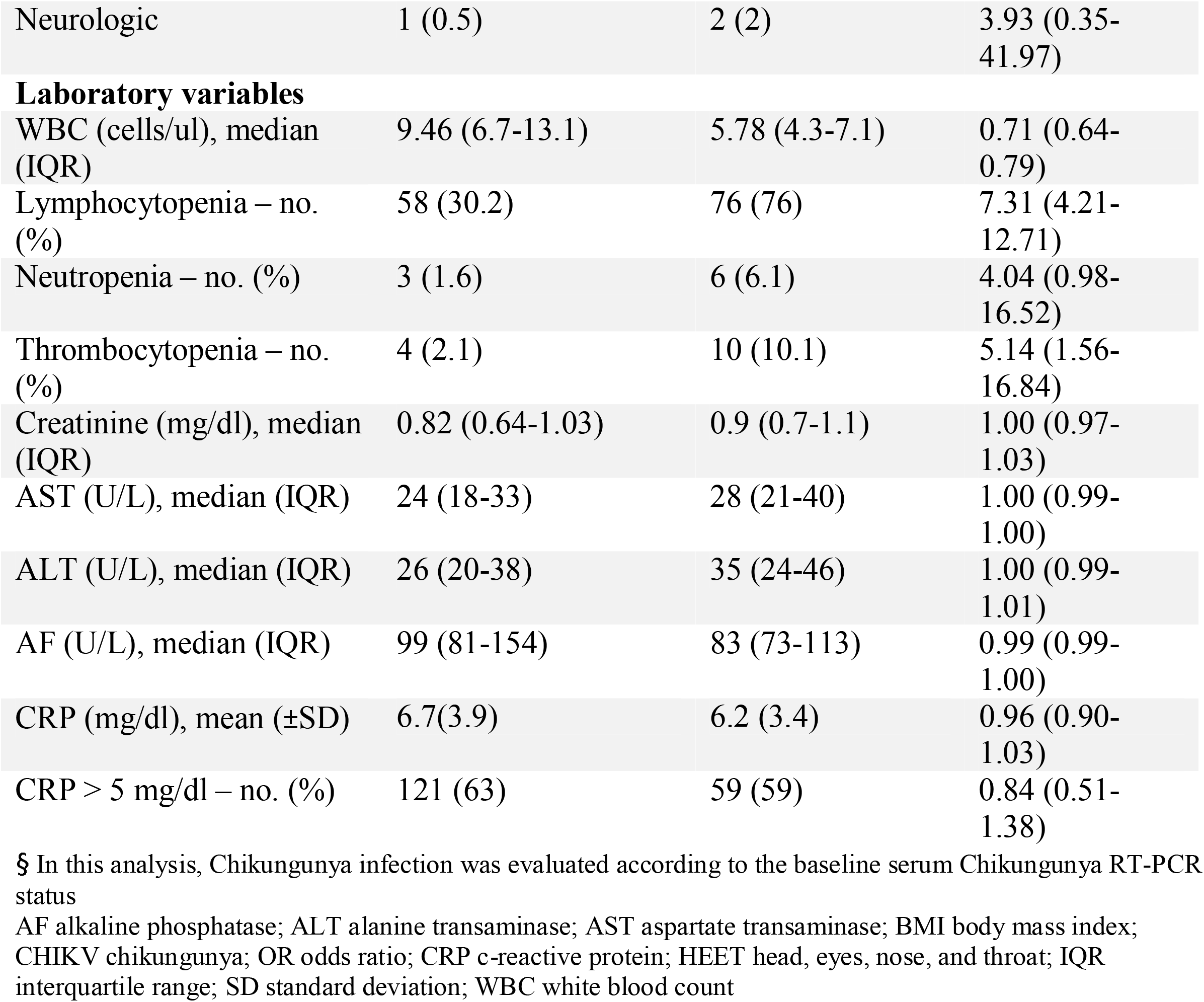
Demographic, clinical and laboratory characteristics of non-severe febrile participants according to chikungunya infection, Rio de Janeiro, Brazil, October 2018–July 2019

**Supplementary Table 3.** Diagnostic performance of the convalescent-phase febrile samples for IgM, IgG and the combination of both IgM and IgG components of the DPP® ZDC IgM/IgG test (Bio-Manguinhos, Fiocruz, Brazil) against Chikungunya RT-PCR, among non-severe febrile patients, Rio de Janeiro, Brazil, October 2018-July 2019.

| Index test | N,<br>sensitivity<br>(95% CI) | N,<br>specificity<br>(95% CI) | PPV<br>(95% CI) | NPV<br>(95% CI) | PLR<br>(95%<br>CI) | NLR<br>(95%<br>CI) | AUC<br>(95%<br>CI) |
| --- | --- | --- | --- | --- | --- | --- | --- |
| **DPP IgM** | 65.75 (53.72-76.47) | 98.11 (89.93-99.95) | 97.96<br>(89.15-99.95) | 67.53<br>(55.90-77.77) | 34.85<br>(4.97-244.57) | 0.35<br>(0.25-0.48) | 79.37<br>(71.25-86.06) |
| **DPP IgG** | 77.78 (64.40-87.96) | 93.33 (85.12-97.80) | 89.36<br>(76.90-96.45) | 85.37<br>(75.83-92.20) | 11.67<br>(4.94-27.54) | 0.24<br>(0.14-0.39) | 86.82<br>(79.74-92.13) |
| **DPP**<br>**IgM/IgG** | 65.75 (53.72-76.47) | 93.42 (85.31-97.83) | 90.57<br>(79.34-96.87) | 73.96<br>(64.00-82.38) | 9.99<br>(4.22-23.70) | 0.37<br>(0.27-0.51) | 79.87<br>(72.52-85.98) |

AUC stands for area under the curve; CI confidence intervals; NPV negative predictive value; NLR negative likelihood ratio; PPV positive predictive value; PLR positive likelihood ration.

## Supplementary Figures

**Supplementary Figure 1.**
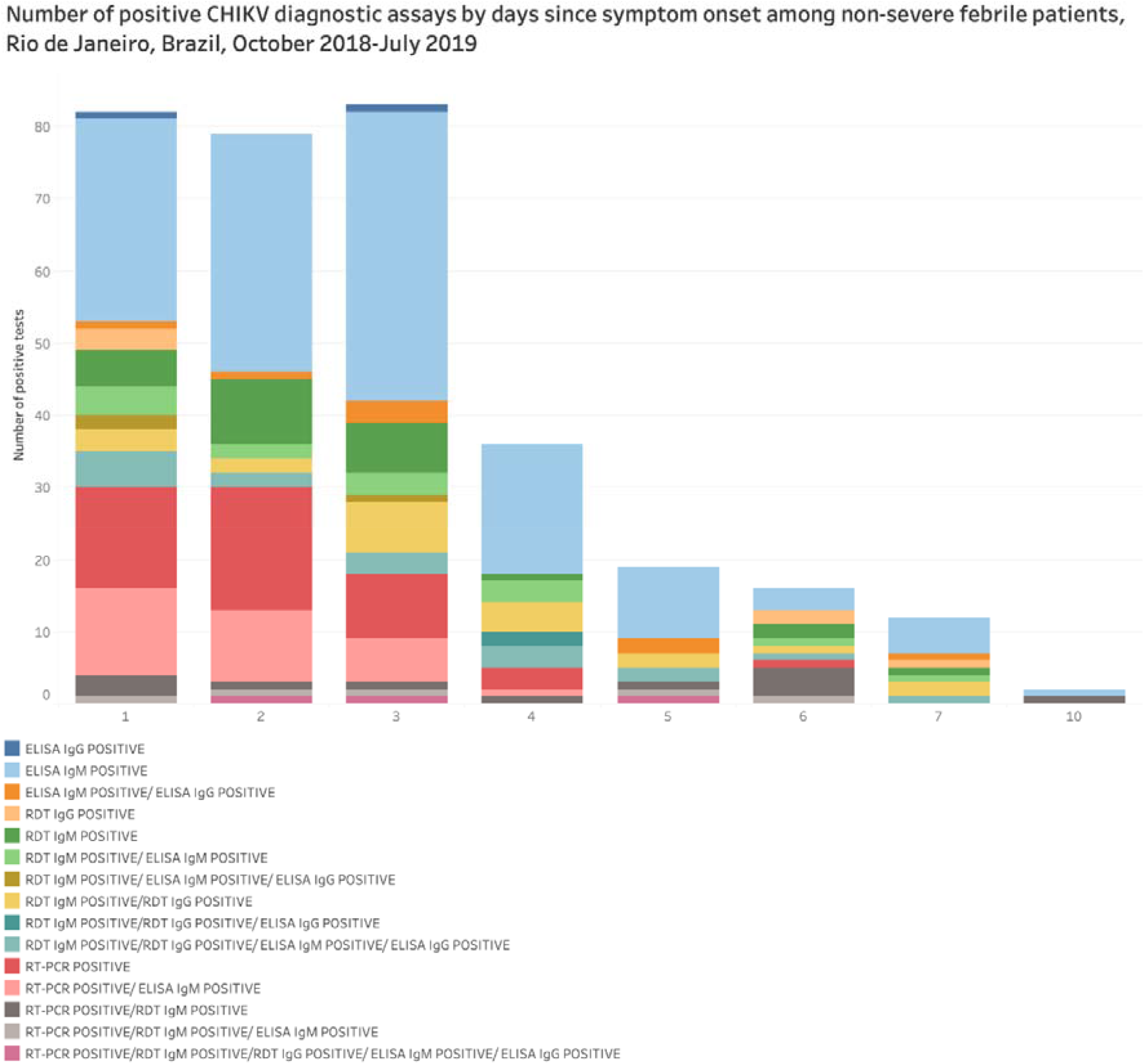
Number of positive Chikungunya diagnostic assays by days since symptom onset among non-severe febrile patients, Rio de Janeiro, Brazil, October 2018-July 2019.

**Supplementary Figure 2.**
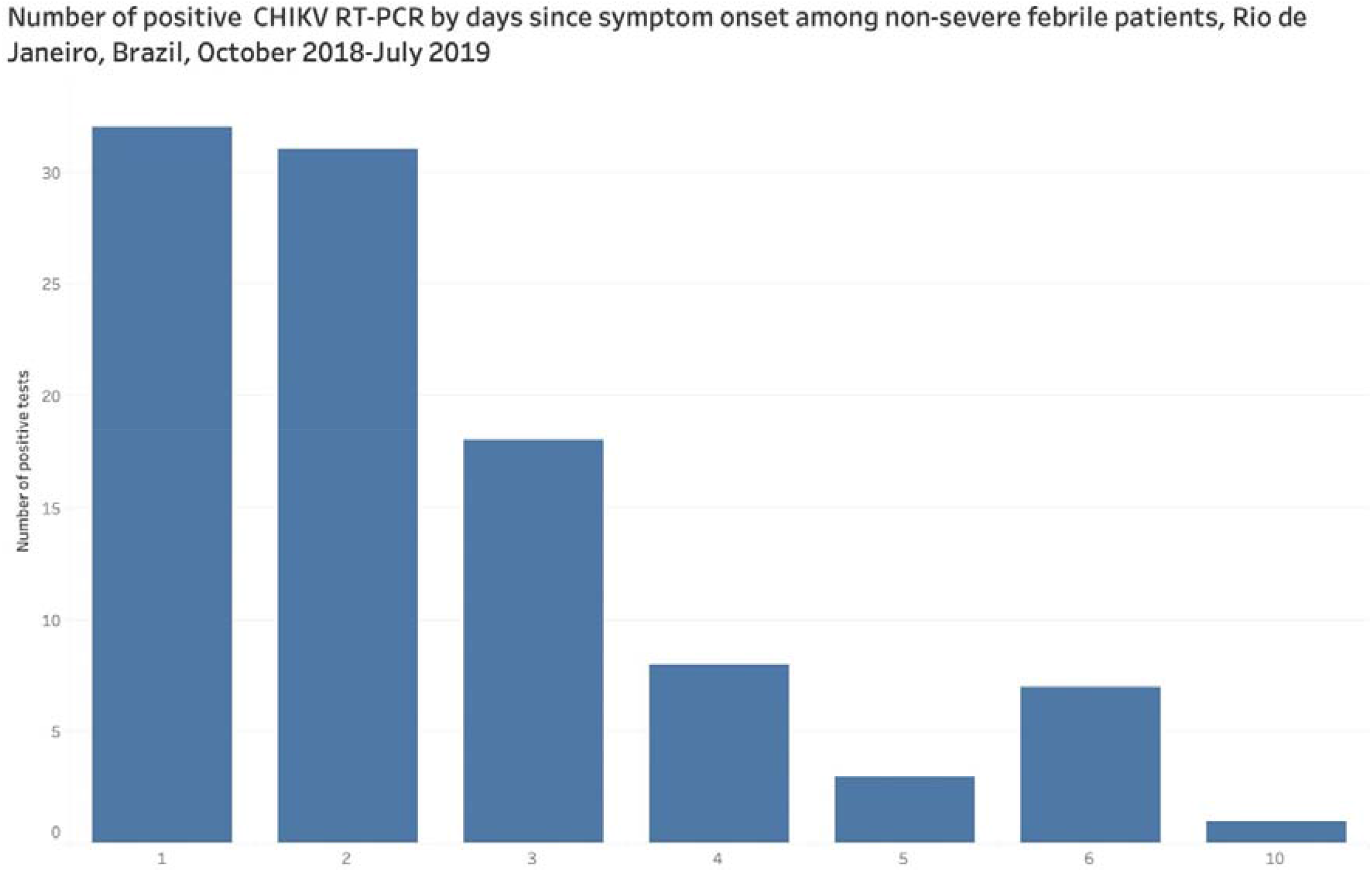
Number of positive Chikungunya reverse-transcriptase polymerase chain reaction by days since symptom onset among non-severe febrile patients, Rio de Janeiro, Brazil, October 2018-July 2019

**Supplementary Figure 3.**
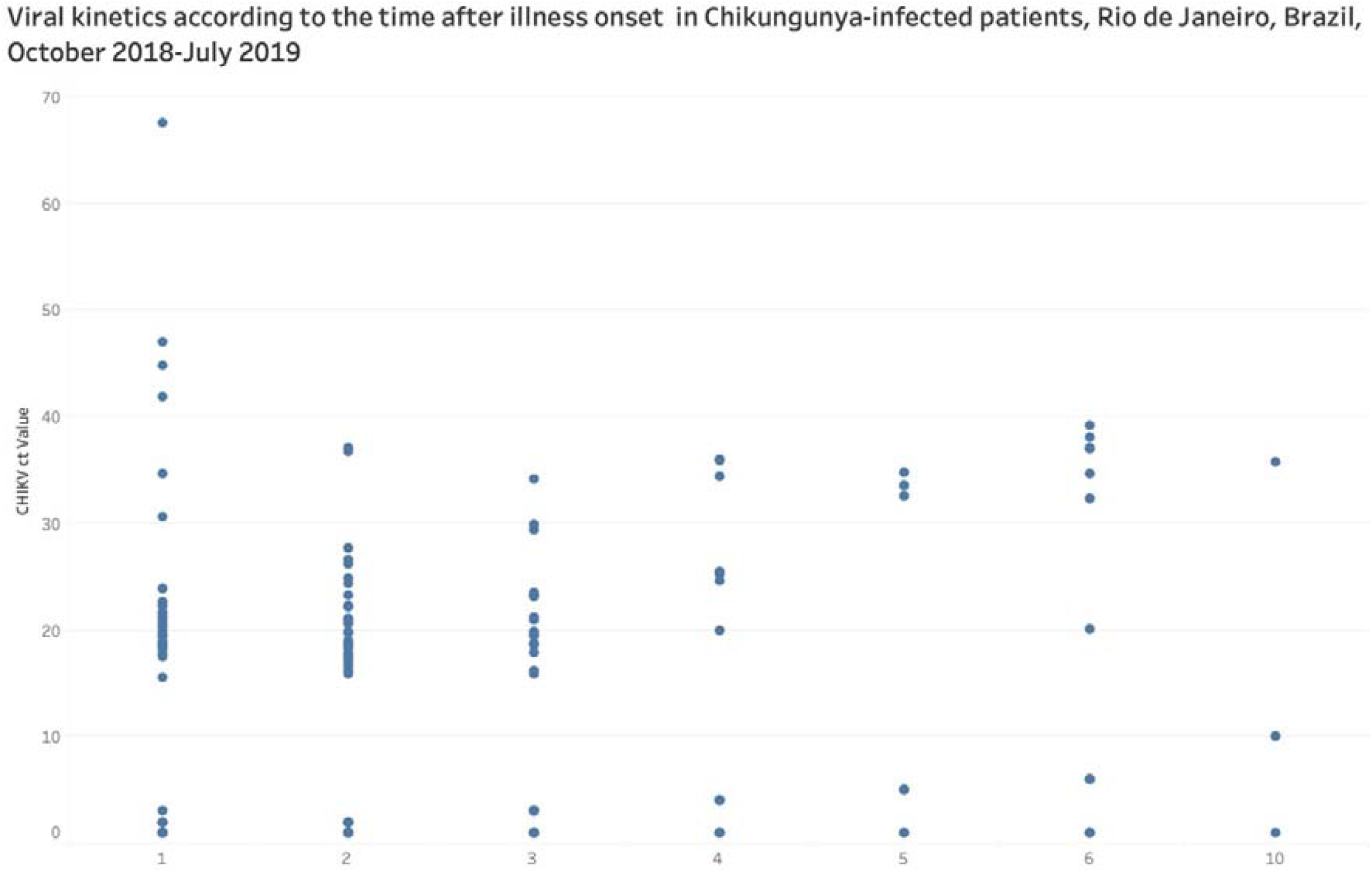
Viral kinetics according to the time after illness onset in Chikungunya-infected patients, Rio de Janeiro, Brazil, October 2018-July 2019

**Supplementary Figure 4.**
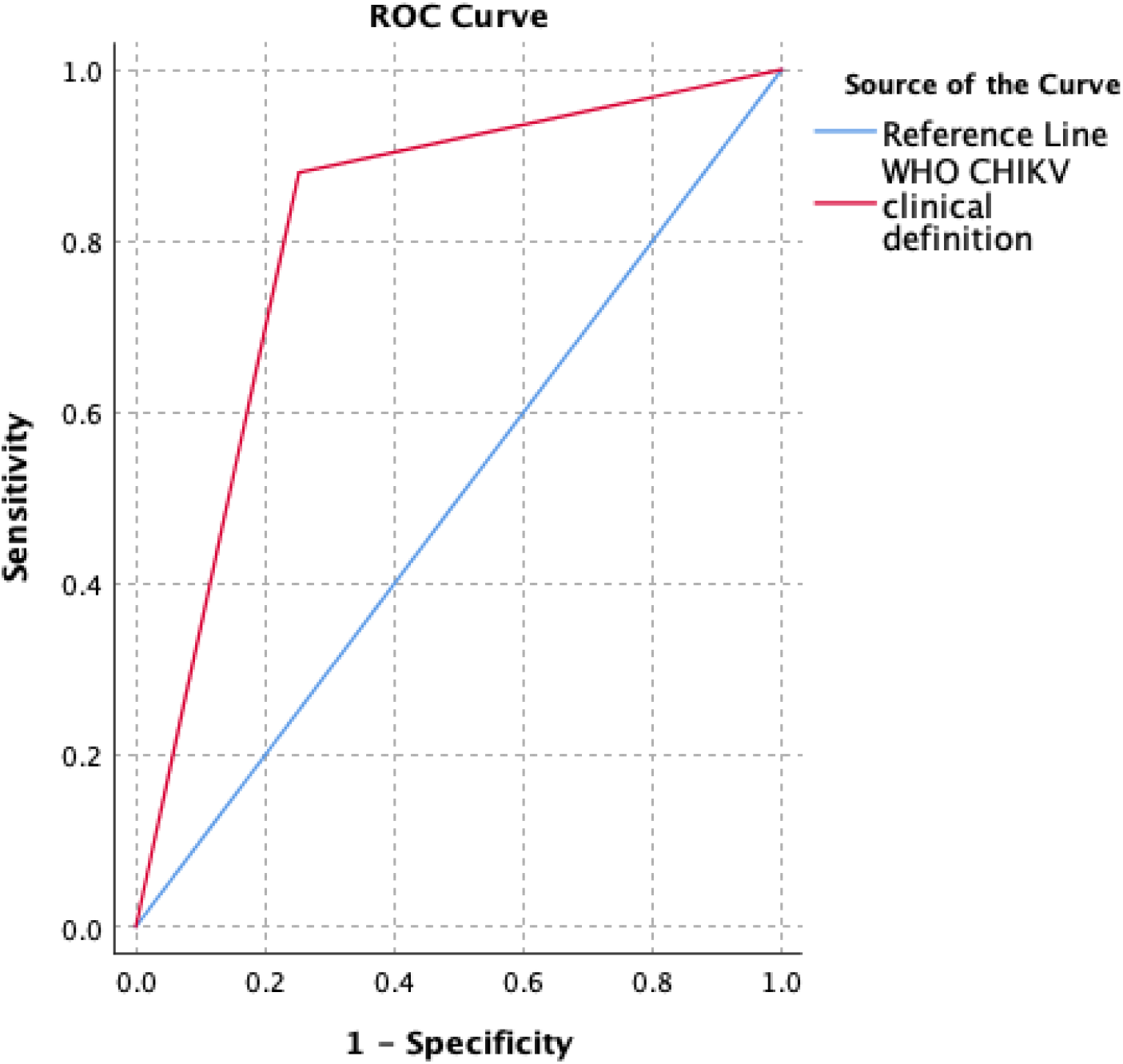
Diagnostic accuracy of the current Chikungunya clinical case definition endorsed by the World Health Organization among non-severe febrile illness patients, Rio de Janeiro, October 2018-July 2019 CHIKV chikungunya virus; WHO World Health Organization; ROC receiver operating curve The clinical definition of CHIKV had a sensitivity of 88% (95% CI: 79.9-93.6) and a specificity of 74.7% (95% CI: 68-80). This definition allowed to classify correctly 81% (95% CI: 76-86) of individuals with non-severe febrile illness who seek care at the primary care in Rio de Janeiro, Brazil.

## Notes

### Clinical Trial

Clinicaltrials.gov: NCT03047642

### Funding Statement

This work was funded by independent grants from Australia, the Netherlands, and the UK aid from the British people.

### Author Declarations

The Brazilian National Ethics Research Committee (CONEP) approved the study (protocol number: 70984617.9.0000.5262), and written informed consent was obtained from all participants or the caregivers of participants before enrollment.

